# Safety and Compliance among Newly Qualified Paramedics in a Pre-hospital Clinical Trial of an Investigational Medicinal Product: A Post-Hoc analysis of the PACKMaN Randomised Controlled Trial

**DOI:** 10.1101/2025.07.31.25332526

**Authors:** A. Rosser, I. Gunson, Z. Green, R. Lall, F. Michelet, E. Miller, J. Miller, H. Noordali, GD. Perkins, O. Stanley, MA. Smyth

## Abstract

**Background:** Pre-hospital research has unique challenges. Ambulance clinicians are required to enrol patients in emergency situations, often remote from the research team at time of recruitment. With Newly Qualified Paramedics (NQPs) representing a significant and growing proportion of ambulance staff, it is important to establish if they can safely and effectively recruit patients to clinical trials. This paper reports a post-hoc analysis of the PACKMaN trial, a large, double-blind Randomised Controlled Trial of an Investigational Medicinal Product (CTIMP), of ketamine versus morphine in the pre-hospital setting.

**Methods:** Adverse Events (AEs) and Serious Adverse Events (SAEs) experienced by patients recruited to the PACKMaN trial, and protocol Non-Compliances (NCs) by paramedics during the trial were retrospectively analysed. We compared recruitment, incidence and type of AEs, as well as incidence of SAEs and NCs dichotomised by paramedic experience.

**Results:** Of the 458 patients, 259 (56.6%) and 199 (43.4%) were recruited by experienced paramedics and NQPs respectively. Incidence of AEs was similar regardless of experience: experienced paramedics reported 128/259 (49.8%) and NQPs reported 91/199 (45.7%) OR 0.86 95% CI [0.60 to 1.25]. SAEs slightly increased, but not statistically significantly, in the NQP group: experienced paramedics 4/259 (1.5%), NQPs 8/199 (4.0%) OR 2.67 95% CI [0.79 to 9.00]. NC was similar amongst both groups, experienced paramedics 3/259 (1.2%), NQPs 6/199 (3.0%), OR 2.65 95% CI [0.66 to 10.74].

**Conclusion:** In a double-blind CTIMP, there was no statistical difference in the incidence of AEs or NCs between NQPs and experienced paramedics. NQPs made an important contribution to patient recruitment in this study, improving the generalisability. SAEs and NCs were rare, and patients received analgesics safely. There was no correlation between experience and AE likelihood, and no safety concerns identified arising from NQP participation. Our findings demonstrate that NQPs can safely recruit patients to clinical trials.

## Background

Pre-hospital care has evolved from an emergency transport system to one where appropriately qualified healthcare professionals use their knowledge and skills to deliver high quality, evidence-based healthcare (Cimino & Braun, 2023; Lowthian et al., 2011; Williams et al., 2009). Pre-hospital clinicians routinely provide the first clinician contact in many healthcare systems and they play a vital role in helping to reduce morbidity and mortality. In the United Kingdom (UK), pre-hospital care is led by paramedics who are autonomous, professionally registered clinicians, with a title protected in law (Eaton, 2023). The role of the paramedic has evolved significantly since its inception. Today, it is less well-defined and better described as a domain of practice that includes a wide range of skills, knowledge and contribution to research (Williams et al., 2021).

To qualify as a paramedic in the UK, one must first complete a 3-year undergraduate degree programme accredited by the Health and Care Professions Council (HCPC). Prior to 2016, the graduate paramedic would have progressed immediately towards unsupervised lead clinician role (The NHS Staff Council, 2017). However, today graduate paramedics must undertake a structured Newly Qualified Paramedic (NQP) programme (NHS Employers, 2019). The NQP programme provides a two-year post qualification foundation period to consolidate skills and knowledge, during which scope of practice may be curtailed, supervised or subject to specific practice restrictions (East of England Ambulance Service, 2024; London Ambulance Service, 2025). It requires that the NQP receives regular supervision and support from senior paramedics and must complete a clinical practice portfolio, including reflective practice and participation in regular scheduled appraisals (College of Paramedics, 2021). This supports the NQP to develop their professional, social and personal identity as a registered paramedic (Phillips & Trenoweth, 2023). Following successful completion of the NQP programme, paramedics are considered sufficiently experienced to no longer require routinely supervised practice.

A large proportion of the NHS ambulance paramedic workforce is comprised of NQPs, accounting for approximately 20% of the total paramedic workforce at both Yorkshire Ambulance Service NHS Trust (YAS) and West Midlands Ambulance Service University NHS Foundation Trust (WMAS). However, in routine frontline clinical practice, both ambulance services report that a quarter of operational paramedics providing face-to-face contact with patients were NQPs. This may have important implications for the delivery of clinical trials within ambulance services.

It has been reported that there were fewer high quality randomised controlled trials (RCTs) completed in ambulance services compared to other healthcare settings and professions (Björklund et al., 2021). This may be attributable to the research culture within ambulance services (Lawrie et al., 2023) and/or the challenging environmental conditions which reduce the required bandwidth for research activity (Cimino & Braun, 2023). This latter point highlights a potential risk relating to adherence to study protocols and concern for patient safety. Current evidence is sparse relating to the participation of research by NQPs and the cognitive burden this might create. How this may impact research participation by NQPs is unclear. Evidence is currently lacking however, there is a plausible risk that inexperienced paramedics may experience bandwidth overload when simultaneously managing clinical and research activity (Copson et al., 2024).

This paper aims to explore recruitment, incidence of Adverse Events (AEs), Serious Adverse Events (SAEs) and Protocol Non-Compliances (NCs), in the PACKMaN trial, dichotomised by paramedic experience. The Paramedic Analgesia Comparing Ketamine and MorphiNe in trauma (PACKMaN) trial was a multi-centre, double-blinded RCT of controlled drugs conducted in two regional NHS ambulance services (Smyth, 2025)

## Methods

PACKMaN compared the clinical and cost effectiveness of ketamine and morphine for severe pain following acute traumatic injury (EudraCT 2020-000154-10, ISRCTN14124474) and was funded by the National Institute for Health and Care Research Health Technology Assessment (National Institute for Health and Care Research, 2024). All study participants were recruited during the period 10/11/2021-16/05/2023.

We conducted a post hoc analysis of the work experience of paramedics recruiting participants into the PACKMaN trial. This analysis was dichotomised by paramedic experience, considered as experienced paramedic (more than 2 years post qualification) or NQP (less than 2 years). We undertook descriptive analyses of trained paramedics, the patients they enrolled, adverse events, serious adverse events (SAEs), and protocol non-compliances for each participating ambulance service relating to the PACKMaN trial recruitment. Chi-square of independence was used to determine the association between variables.

This post hoc analysis was conducted of the PACKMaN trial data, without *a priori* intent within the study protocol, and expands upon a midpoint analysis requested by the Trial Steering Committee. This analysis was requested to provide assurance of safety with NQP participation, during the participant recruitment phase of the trial. This post hoc analysis includes all 458 patient participants and all 489 paramedic participants.

### Setting

PACKMaN was undertaken in West Midlands Ambulance Service University NHS Foundation Trust (WMAS) and Yorkshire Ambulance Service NHS Trust (YAS). WMAS serves a population of 5.6 million over a geographical area of 5500 square miles (West Midlands Ambulance Service, 2024) while YAS serve 5 million people over 6000 square miles (Yorkshire Ambulance Service, 2024). Collectively, these services receive more than 2 million 999 emergency calls annually and employ more than 14,000 staff. WMAS employs more than 2700 paramedics, with 88% of these routinely providing clinical response to 999 calls. Of these paramedics, 25.2% are NQPs. YAS employs almost 2200 paramedics, of whom 22.8% are NQPs routinely providing clinical response to 999 calls.

### Participants

All registered paramedics at participating ambulance stations, regardless of prior experience, were eligible to participate in the PACKMaN trial. We dichotomised participating paramedics into NQP and experienced groups. At the time of undertaking training for PACKMaN, paramedics who were undertaking their NQP phase were categorised as NQP, while paramedics who had successfully completed their NQP phase were categorised as experienced.

All participating paramedics, irrespective of experience received a bespoke training package to ensure understanding of the trial procedures, pharmacology of trial medicines and outcome reporting. Training was delivered either through online or face to face delivery. All participating paramedics were required to complete a Patient Group Directive to supply or administer the IMP and Midazolam, in accordance with the study protocol.

### Patient and Public Involvement

Patient and Public Involvement (PPI) was embedded into the PACKMaN trial throughout. PPI featured at all stages, through design, study materials, conduct and dissemination. We obtained broad representation through those with personal experience of severe trauma and specialist PPI groups.

### Outcomes

During the PACKMaN trial all AEs, SAEs and NCs were recorded, analysed and findings reported to the independent Data Monitoring Committee (DMC). We linked each of these events to the participating paramedic to assess if their prior level of experience might have been a contributing factor.

AEs and SAEs were pre-specified, well-defined, and were reported by recruiting clinicians, hospital clinicians or study research teams as soon as they were identified. Safety reporters were blinded to the study arm and to the paramedic group (NQP or experienced).

An AE was defined as any untoward medical occurrence in a patient administered a medicinal product. Predictable drug side effects were recorded as AEs. An SAE is an AE that fulfils one or more of the following criteria:

- Results in death
- Is immediately life-threatening
- Requires hospitalisation or prolongation of existing hospitalisation
- Results in persistent or significant disability or incapacity
- Causes a congenital abnormality or birth defect
- Requires immediate intervention to prevent one of the above or is otherwise an important medical condition.

Non-compliance was defined as deviation from the trial protocol or trial related procedures. Non-compliance was not limited to clinician-patient interaction. For example, failure to complete case report forms could be a non-compliance event. In this analysis we have reported NCs attributable to participating paramedics only, we have not reported NCs attributable to other causes e.g. trial team staff.

We report descriptive statistics, Fisher’s exact test, chi-square and logistic regression to explore the incidence and significance (p=<0.05) of recruitment, AEs, SAEs and NCs dichotomised by paramedic experience (NQP vs experienced paramedic).

## Results

In total 489 paramedics completed the training necessary to recruit patients to PACKMaN. Participating paramedics were more likely to be experienced paramedics (66.9%), than NQPs (33.1%). There was no significant difference in participation between the two ambulance services (Table 1). During the study, experienced paramedics recruited 56.6% of patients while NQPs recruited 43.4%. NQPs in YAS recruited greater proportion of patients than NQPs in WMAS (Table 1).

**Table 1:** Experience and recruitment by participating paramedics.

| **Number of paramedics<sup>1</sup>** | **WMAS n (%)** | **YAS n (%)** | **Total n (%)** |
| --- | --- | --- | --- |
| Experienced paramedic | 171 (69.5) | 156 (64.2) | 327 (66.9) |
| NQP | 75 (30.5) | 87 (35.8) | 162 (33.1) |
| **Recruitment by paramedic<sup>2</sup>** |  |  |  |
| Experienced paramedic | 149 (64.0) | 110 (48.9) | 259 (56.6) |
| NQP | 84 (36.0) | 115 (51.1) | 199 (43.4) |

<sup>1</sup> chi square (1, $N = 489$ ) = 1.5586, $p = 0.212$
<sup>2</sup> chi square (1, $N = 458$ ) = 10.5652, $p = 0.001$

PACKMaN employed a double-blind randomisation strategy to conceal the medication administered from both paramedics and patients. Our analysis by treatment arms indicates that there was no significant difference in the number of patients recruited to each arm of the study across paramedic experience levels at either site, nor was there any difference in the study arm randomisation according to experience level (Table 2).

**Table 2:** Recruitment by trial arm.

| **Recruitment by service<sup>1</sup>** | **Morphine n (%)** | **Ketamine n (%)** | **Total n (%)** |
| --- | --- | --- | --- |
| YAS | 113 (48%) | 112 (50%) | 225 |
| WMAS | 121 (52%) | 112 (50%) | 233 |
| **Recruitment by experience<sup>2</sup>** |  |  |  |
| NQP | 106 (45.3%) | 93 (41.5%) | 199 (43.4%) |
| Experienced paramedic | 128 (54.7%) | 131 (58.5%) | 259 (56.6%) |

<sup>1</sup> chi square (1, $N = 458$ ) = 0.1338, $p = 0.715$
<sup>2</sup> chi square (1, $N = 458$ ) = 0.666, $p = 0.414$

Throughout the study period, 219 patients (47.8%) experienced at least one AE, 12 patients (2.6%) suffered SAEs and there were 10 NC events involving 9 patients (2.0%) (Table 3). Some patients experienced more than one adverse event simultaneously, for example, experiencing both respiratory depression and hypotension at the same time. Cumulative reported adverse events by clinical category are reported in Table 4. The vast majority of AEs were clinically predictable side effects of morphine or ketamine. There was no difference in occurrence of AEs amongst paramedics attributable to level of experience [OR 0.86 (0.60 to 1.25); p=0.43].

**Table 3:** Safety related events by paramedic experience level.

| Safety events by experience | Experienced paramedic (n=259) | NQP (n=199) | Unadjusted Odds Ratio (OR) (95% CI); p-value |
| --- | --- | --- | --- |
| Adverse Events n (%) | 128 (49.4%) | 91 (45.7%) | OR 0.86 (0.60 to 1.25); 0.43 |
| Serious Adverse Events n (%) | 4 (1.5%) | 8 (4.0%) | OR 2.67 (0.79 to 9.00); 0.10 |
| Non-compliances n (%) | 3 (1.2%) | 6 (3.0%) | OR 2.65 (0.66 to 10.74); 0.16 |

**Table 4:** Number of adverse events by category split by trial arm and paramedic experience level.

|  | Trial arm | Airway (n) | Respiratory (n) | Cardiovascular (n) | Neurologic (n) | Other (n) |
| --- | --- | --- | --- | --- | --- | --- |
| NQP | Morphine | 13 | 18 | 22 | 10 | 24 |
| Experienced paramedic | Morphine | 14 | 20 | 12 | 8 | 33 |
| NQP | Ketamine | 9 | 7 | 10 | 18 | 14 |
| Experienced paramedic | Ketamine | 7 | 8 | 16 | 24 | 20 |

Among the 12 SAEs, 4 occurred amongst experienced paramedics and 8 amongst NQPs (Table 5). Despite the apparent increased frequency of SAE among NQPs, the difference was not statistically significant [OR 2.67 (0.79 to 9.00); p=0.10]. Of the 10 NC events, 3 were related to patient recruitment by experienced paramedics and 6 related to patient recruitment by NQPs (Table 3). Again, this difference was not statistically significant [OR 2.65 (0.66 to 10.74); p=0.16].

**Table 5:** Serious Adverse Events and Non-Compliance.

| Serious Adverse Events by paramedic experience<sup>1</sup> | Morphine (n) | Ketamine (n) | Total (n) |
| --- | --- | --- | --- |
| NQP | 6 | 2 | 8 |
| Experienced paramedic | 2 | 2 | 4 |
| Non-compliance by paramedic experience<sup>2</sup> |  |  |  |
| NQP | 5 | 2 | 7 |
| Experienced paramedic | 2 | 1 | 3 |

<sup>1</sup> chi square (1, $N = 12$ ) = 0.7500, $p = 0.386$
<sup>2</sup> chi square (1, $N = 10$ ) = 0.0227, $p = 0.880$

Similarly, when we examined SAEs and NCs by trial arm we failed to identify any statistically significant difference attributable to level of experience (Table 5).

## Discussion

In this analysis of data from the PACKMaN trial we found that NQPs did not experience a higher incidence of AEs, SAEs or NCs compared to their more experienced paramedic colleagues. This suggests that participation of NQPs in clinical research does not increase the risk of safety related events for patients enrolled in clinical trials. We found no evidence of safety implications to including NQPs in this trial. Consequently, NQPs should not be discouraged from participating in research on patient safety grounds.

Our analysis is supported by findings from other clinical trials conducted in UK Ambulance Services. Clinical trials of devices including PARAMEDIC (Perkins et al., 2015), AIRWAYS-2 (Benger et al., 2018) and ACUTE (Fuller et al., 2020) and Clinical Trials of Investigational Medicinal Products (CTIMP) such as PARAMEDIC2 (Perkins et al., 2018) and RIGHT-2 (RIGHT-2 Investigators, 2019) have permitted participation by NQPs and reported low incidence of AEs and SAEs.

In the PACKMaN trial approximately one-third of participating paramedics were NQPs. This proportion is broadly reflective of the experience mix seen in clinical practice across ambulance services more generally. The inclusion of NQPs in the PACKMaN trial also reflects the pragmatic nature of ambulance services both with respect to participation in research and the introduction of new medicines. NQPs comprise a significant proportion of the operational workforce. Inclusion of NQPs in clinical research is therefore more representative of workforce capability and findings will therefore be more generalisable. Furthermore, if ketamine is added to the paramedic formulary, then NQPs may be expected to use it safely. Inclusion of NQPs in PACKMaN has helped to establish that ketamine can be used safely by NQPs.

We recognise that the paramedics who volunteered to participate in the PACKMaN trial may not truly be representative of the wider paramedic workforce, and we did not record the motivating reason why some paramedics choose to participate. None the less, we are hopeful that these research findings would be generalisable to the wider paramedic workforce, including the NQP workforce. Furthermore, we would caution that findings from clinical research studies that exclude NQPs may not be generalisable to the UK ambulance service clinical workforce.

It is also important to acknowledge that NQPs may play a vital role in recruitment to clinical trials. In PACKMaN, NQPs were responsible for recruiting 43.4% of participants. Without NQP participation, recruitment would have taken considerably longer, incurring substantial and avoidable costs. Excluding NQPs from participation in clinical trials without robust justification may render clinical trials in ambulance services financially unviable. Failure to undertake research may unintentionally extend the provision of sub-optimal treatments, leading to increased harm or suffering.

It has been suggested that when managing complex or severely injured patients, NQPs may experience high levels of stress and be close to being overwhelmed. In such cases, it may be inappropriate to expect NQPs to undertake research activities, when they might be struggling to manage a complex clinical case. This argument asserts that NQPs have limited bandwidth and that research activities consume scarce bandwidth, potentially to the detriment of both the NQP and to patient care (Lawrie et al., 2023). We believe our findings refute this argument; we found no evidence of safety implications to including NQPs in this trial. However, we acknowledge that these claims must not be overstated, and in the presence of sparse and potentially conflicting evidence it is clear that further research is required, exploring the impacts of cognitive load amongst NQPs.

Although ketamine is used routinely by non-specialist paramedics in Australia, New Zealand, South Africa, the USA, Canada and several European countries, in the UK its use has been restricted to critical care and specialist paramedics where its primary indications include analgesia for severe pain unresponsive to morphine and procedural sedation. The inclusion of ketamine for non-specialist paramedics was always likely to provoke emotive views amongst some communities, further heightened by the involvement of NQPs. Some services mandate that ketamine administration requires anaesthetic level monitoring, and clinicians must have advanced airway management capability (Aldridge et al., 2023; Morgan et al., 2021; West Midlands Ambulance Service, 2021).

In the PACKMaN trial, patients received routine non-invasive physiologic monitoring, paramedics did not use end tidal carbon dioxide (EtCO2) monitoring or require the presence of clinicians competent to perform endotracheal intubation (ETI). Despite a lower level of physiologic monitoring and lack of ETI capability, patients did not experience an increased incidence of AEs (Smyth et al., 2025). Furthermore, as we have already reported there was no significant difference in AEs attributable to experience. Ketamine therefore appears to be safe in the hands of NQPs providing sub-dissociative pain relief. Excluding NQPs from research participation would have limited the generalisability of the study to UK practice, where NQPs are an important group of the ambulance workforce.

Finally, even though NQPs are undertaking a development pathway, as HCPC registrants they are still accountable to the professional body and held to the same standard as their fellow registrants. Consequently, there is a professional expectation that NQPs are able to provide the same standard of care expected of a more experienced paramedic. We are grateful to our NQPs who took part in this study. They made an important contribution, both in terms of patient enrolment, but also in improving the generalisability of study findings across the ambulance setting. We recommend the default approach in pre-hospital research should be to include NQPs

## Limitations

Our study has four notable limitations. First, the study required NQPs and paramedics to volunteer to participate in the PACKMaN trial. The reason why individual NQPs or experienced paramedics opted to participate, or more importantly not participate in the study was not captured. We did not record their confidence in managing patients with traumatic injury, confidence in delivering potent analgesics or confidence to participate in a CTIMP. We did not measure the cognitive impact of NQPs and paramedics recruiting patients to a controlled drug CTIMP. It is therefore unclear, how these study findings relate to routine NQP and paramedic practice amongst a wider workforce and provides opportunity for further research to understand those cognitive loads.

Second, the data presented formed a post-hoc analysis of the PACKMaN study data, we did not precisely capture the experience of participating paramedics. For example, an NQP who had attended university straight from school was considered the same as an NQP who had 10 years of experience as an ambulance technician prior to attending university. Similarly, we dichotomised paramedics at the time they completed trial related training. Paramedics who transitioned from NQP, to experienced status, after they had completed trial training were still classified as NQP. We did not collect any detail relating to progress in the NQP programme and are therefore unable to describe how NQPs were distributed across the two-year period.

Third, PACKMaN was conducted in WMAS and YAS only, therefore, the generalisability to NQP participation in research amongst other UK ambulance services is not certain. We are buoyed by infrequent rates of adverse event and protocol non-compliance within two large regional ambulance services and saw no reason why a larger cohort of participating paramedics might adversely impact their occurrence.

Fourth, the findings of this study may not be transferable to international settings due to training, education, and professional differences to UK paramedics.

## Conclusion

The incidence of AEs, SAEs and NC in PACKMaN, a CTIMP of controlled drugs, was low. We found no evidence to suggest that experience of the participating paramedic was a potential contributing factor among AEs, SAEs or NCs. Our findings further demonstrate that NQPs recruited almost half of all patients participating in the PACKMaN trial, suggesting NQPs may have a vital role to play in ensuring clinical trials recruit to target in a timely manner. Finally, because NQPs comprise around one third of the operational workforce in UK Ambulance Services, inclusion of NQPs in research will significantly improve generalisability of research findings to real world clinical practice. We recommend that NQPs should be encouraged to participate in research.

## Data Availability

All data produced in the present work are contained in the manuscript. Reporting of the wider PACKMaN study is available https://doi.org/10.1016/j.lanepe.2025.101265

https://www.thelancet.com/journals/lanepe/article/PIIS2666-7762(25)00057-2/fulltext

## Ethics Approval Statement

West of Scotland REC 1, reference 20/WS/0126, approval granted 01/09/2020. Written informed consent was obtained from all participants, according to the study protocol (Michelet et al., 2023).

## Author Contributions

All authors made substantial contribution to the data processing, interpretation and production of the article. Their contribution extended to the reviewing and revising the article, to ensure accuracy, MS conceived and was the co-chief investigator with GP of the PACKMaN trial. MS and HN managed the PACKMaN trial centrally. FM provided data analysis and statistical support, with oversight from RL. AR, IG, JM managed the study at WMAS. OS, ZG, EM and JM contributed to data collection. EM managed the study at YAS.

Artificial Intelligence was not used anytime during the analysis, interpretation and production of the article.

## References

Aldridge, J., Quinn, S. A., Oh, S., & Trisler, D. (2023). Ketamine for Pre-Hospital Analgesia and Sedation in the Trauma Population: a Narrative Review. SN Comprehensive Clinical Medicine, 5(1), 63. 10.1007/s42399-023-01409-z

Benger, J. R., Kirby, K., Black, S., Brett, S. J., Clout, M., Lazaroo, M. J., Nolan, J. P., Reeves, B. C., Robinson, M., Scott, L. J., Smartt, H., South, A., Stokes, E. A., Taylor, J., Thomas, M., Voss, S., Wordsworth, S., & Rogers, C. A. (2018). Effect of a Strategy of a Supraglottic Airway Device vs Tracheal Intubation During Out-of-Hospital Cardiac Arrest on Functional Outcome: The AIRWAYS-2 Randomized Clinical Trial. Jama, 320(8), 779–791. 10.1001/jama.2018.11597

Björklund, M. K., Cruickshank, M., Lendrum, R. A., & Gillies, K. (2021). Randomised controlled trials in pre-hospital trauma: a systematic mapping review. Scand J Trauma Resusc Emerg Med, 29(1), 65. 10.1186/s13049-021-00880-8

Cimino, J., & Braun, C. (2023). Clinical Research in Prehospital Care: Current and Future Challenges. Clin Pract, 13(5), 1266–1285. 10.3390/clinpract13050114

College of Paramedics. (2021). Scope of Practice. College of Paramedics. Retrieved 10/12/2024 fro https://collegeofparamedics.co.uk/COP/ProfessionalDevelopment/Scope_of_Practice.aspx

Copson, J., Eaton, G., & Mahtani, K. R. (2024). Transition processes for newly qualified paramedics entering primary care: a critical discussion and theoretical perspective. Br J Gen Pract, 74(742), 228–231. 10.3399/bjgp24x737337

East of England Ambulance Service. (2024). Scope of Practice Policy - POL092. East of England Ambulance Service NHS Trust Retrieved from https://content.eastamb.nhs.uk/assets/Scope_of_Practice_Policy_84cfabb7f9.pdf

Eaton, G. (2023). Addressing the challenges facing the paramedic profession in the United Kingdom. Br Med Bull, 148(1), 70–78. 10.1093/bmb/ldad024

Fuller, G., Keating, S., Goodacre, S., Herbert, E., Perkins, G., Rosser, A., Gunson, I., Miller, J., Ward, M., Bradburn, M., Thokala, P., Harris, T., Marsh, M., Scott, A., & Cooper, C. (2020). Is a definitive trial of prehospital continuous positive airway pressure versus standard oxygen therapy for acute respiratory failure indicated? The ACUTE pilot randomised controlled trial. BMJ Open, 10(7), e035915. 10.1136/bmjopen-2019-035915

Lawrie, L., Duncan, E. M., Lendrum, R., Lebrec, V., & Gillies, K. (2023). Challenges and opportunities for conducting pre-hospital trauma trials: a behavioural investigation. Trials, 24(1), 157. 10.1186/s13063-023-07184-5

London Ambulance Service. (2025). Newly Qualified Paramedic roles – FAQs. etrieved 30.07.2025 from https://www.londonambulance.nhs.uk/working-for-us/career-opportunities/newly-qualified-paramedic-roles-faqs/

Lowthian, J. A., Cameron, P. A., Stoelwinder, J. U., Curtis, A., Currell, A., Cooke, M. W., & McNeil, J. J. (2011). Increasing utilisation of emergency ambulances. Aust Health Rev, 35(1), 63–69. 10.1071/ah09866

Michelet, F., Smyth, M., Lall, R., Noordali, H., Starr, K., Berridge, L., Yeung, J., Fuller, G., Petrou, S., Walker, A., Mark, J., Canaway, A., Khan, K., & Perkins, G. D. (2023). Randomised controlled trial of analgesia for the management of acute severe pain from traumatic injury: study protocol for the paramedic analgesia comparing ketamine and morphine in trauma (PACKMaN). Scand J Trauma Resusc Emerg Med, 31(1), 84. 10.1186/s13049-023-01146-1

Morgan, M. M., Perina, D. G., Acquisto, N. M., Fallat, M. E., Gallagher, J. M., Brown, K. M., Ho, J., Burnett, A., Lairet, J., Rowe, D., & Gestring, M. L. (2021). Ketamine Use in Prehospital and Hospital Treatment of the Acute Trauma Patient: A Joint Position Statement. Prehosp Emerg Care, 25(4), 588–592. 10.1080/10903127.2020.1801920

National Institute for Health and Care Research. (2024). A randomised controlled trial of Paramedic Analgesia Comparing Ketamine and MorphiNe in trauma (PACKMaN). National Institute for Health and Care Research. Retrieved 13/12/2024 from https://fundingawards.nihr.ac.uk/award/NIHR128086

NHS Employers. (2019). Paramedic consolidation of learning. NHS Employers. https://www.nhsemployers.org/articles/paramedic-consolidation-learning

Perkins, G. D., Ji, C., Deakin, C. D., Quinn, T., Nolan, J. P., Scomparin, C., Regan, S., Long, J., Slowther, A., Pocock, H., Black, J. J. M., Moore, F., Fothergill, R. T., Rees, N., O’Shea, L., Docherty, M., Gunson, I., Han, K., Charlton, K.,… Lall, R. (2018). A Randomized Trial of Epinephrine in Out-of-Hospital Cardiac Arrest. N Engl J Med, 379(8), 711–721. 10.1056/NEJMoa1806842

Perkins, G. D., Lall, R., Quinn, T., Deakin, C. D., Cooke, M. W., Horton, J., Lamb, S. E., Slowther, A. M., Woollard, M., Carson, A., Smyth, M., Whitfield, R., Williams, A., Pocock, H., Black, J. J., Wright, J., Han, K., & Gates, S. (2015). Mechanical versus manual chest compression for out-of-hospital cardiac arrest (PARAMEDIC): a pragmatic, cluster randomised controlled trial. Lancet, 385(9972), 947–955. 10.1016/s0140-6736(14)61886-9

Phillips, P., & Trenoweth, S. (2023). Crossing the ‘flaky bridge’ - the initial transitory experiences of qualifying as a paramedic: a mixed-methods study. Br Paramed J, 8(1), 18–27. 10.29045/14784726.2023.6.8.1.18

RIGHT-2 Investigators. (2019). Prehospital transdermal glyceryl trinitrate in patients with ultra-acute presumed stroke (RIGHT-2): an ambulance-based, randomised, sham-controlled, blinded, phase 3 trial. Lancet, 393(10175), 1009–1020. 10.1016/s0140-6736(19)30194-1

Smyth, M. A., Noordali, H., Starr, K., Yeung, J., Lall, R., Michelet, F., Fuller, G., Petrou, S., Walker, A., Green, Z., McLaren, R., Miller, E., Buckley, D., & Perkins, G. D. (2025). Paramedic analgesia comparing ketamine and morphine in trauma (PACKMaN): a randomised, double-blind, phase 3 trial. The Lancet Regional Health – Europe, 53. 10.1016/j.lanepe.2025.101265

West Midlands Ambulance Service. (2021). Patient Group Direction for Ketamine Use. West Midlands Ambulance Service University NHS Foundation Trust

West Midlands Ambulance Service. (2024). About Us. West Midlands Ambulance Service University NHS Foundation Trust. Retrieved 10/12/2024 from https://www.wmas.nhs.uk/about/

Williams, B., Beovich, B., & Olaussen, A. (2021). The Definition of Paramedicine: An International Delphi Study. J Multidiscip Healthc, 14, 3561–3570. 10.2147/jmdh.S347811

Williams, B., Onsman, A., & Brown, T. (2009). From stretcher-bearer to paramedic: the Australian paramedics’ move towards professionalisation. Australasian Journal of Paramedicine, 7, 1–12.

Yorkshire Ambulance Service. (2024). Yorkshire Ambulance Service NHS Trust. Retrieved 10/12/2024 from https://www.yas.nhs.uk/about-us/about-us/

